# Recruitment, Consent and DNA Sample Acquisition in a U.S. Precision Health Cohort During the COVID-19 Pandemic

**DOI:** 10.1101/2023.09.25.23289158

**Authors:** Allyson M. Derry, Yvette Strong, Davia Schioppo, Joni Cotter, Geisa M. Wilkins, Laura I. Siqueiros, Andrea Ouyang, Kathleen Hulseman, Joseph Petrosino, Lorrin Liang, Megan Stevenson, Tiffany Elianne Aguilera, Alexandria L. Soto, Katherine Meurer, Alison L. Herman, Inessa Cohen, Guido J. Falcone, Erin E. Longbrake, Cassius I. Ochoa Chaar, Kelly M. Anastasio, Michael F. Murray

## Abstract

**Aim:** The Yale Generations Project (YGP) is a precision health cohort initiative that began enrollment in New Haven Connecticut USA in July 2019. In March 2020, after nine months of operation, pandemic restrictions prompted abrupt changes to staff availability as well as changes to the project’s recruitment, consenting, and sample acquisition. This manuscript describes the successful addition of remote recruitment, consenting, and DNA sampling to YGP workflows during the initial 27-months of pandemic restrictions ending June 30, 2022.

**Methods:** The initial YGP protocol established face-to-face workflow for recruiting, consenting and peripheral blood collection. A telemedicine consent protocol was initiated in April of 2020, and a remote saliva collection was established in October of 2020. De-identified data was extracted from YGP dataset and reported here.

**Results:** At the completion of YGP’s initial 36 months (9-months pre-pandemic and 27-months pandemic) YGP enrolled N=4949 volunteers. There were N=1,950 (216.7 per month) volunteers consented pre-pandemic and N=2,999 (111.1 per month) during pandemic. The peak consenting month was February 2020 with N=428. DNA sample acquisition peaked in the pre-pandemic month of February 2020 with N=291 peripheral blood draws, and in the pandemic period the peak DNA acquisition month was November 2020 with N=176 (N=68 peripheral blood draws and N=108 saliva samples).

**Conclusion:** The YGP successfully transitioned from pre-pandemic recruiting, consenting and sample acquisition model that was exclusively face-to-face, to pandemic model that was predominantly remote. The added value of remote recruiting, consenting, and sampling has led to plans for an optimized hybrid model post-pandemic.

## INTRODUCTION

The healthcare infrastructure in the United States includes 637 distinct health systems ^**[1]**^. In the last twenty years, a small number of these health systems have established precision health cohorts in which volunteers are recruited, consented, and provide a DNA sample for genomic sequencing, and are then given genomic screening results that can be used for pharmacogenomic decision making and for preventive care related to identified monogenic risk for predisposition to cancer and heart disease ^**[2-5]**^.

The Yale New Haven Health system (YNHH) together with the Yale School of Medicine established such a program in July of 2019 in the state of Connecticut. This project was named the Yale Generations Project (YGP) which had a formal launch ceremony in September of 2019 ^**[6]**^. The YGP strategy included a pre-launch phase from July 2019 until September 2019, followed by a recruitment ramp-up phase. The recruitment ramp-up phase was expected to last an additional nine months, with the goal of reaching a steady state of enrollment of greater than 800 volunteers per month by the end of year one (i.e., June 2020). The state of Connecticut, which borders New York in the northeastern region of the country, saw the beginning of significant community spread of SARS-CO2 during March of 2020 ^**[7]**^. By the end of that month COVID-19 institutional precautions were undertaken, including suspension of outpatient encounters related to non-therapeutic research at Yale. YGP has observed all institutional and governmental infection control mandates during the COVID-19 pandemic period described in this manuscript, and the effect of these mandates on protocols and growth of the cohort are described.

## METHODS

The YGP plan was to launch the project in the second half of 2019 and to first establish and optimize face-to-face recruitment, consenting and DNA sample collection. There was a plan to explore remote options for recruitment, consenting and DNA collection once: [1] face-to-face workflows were fully optimized, and [2] the project staffing plan of eleven “full-time equivalents” (FTEs) consenters was realized.

During March of 2020, hiring was halted at nine full-time staff, who increasingly needed to be urgently diverted to other projects and duties with rising local COVID-19 cases. In addition to staffing interruptions in March 2020, there were increasing interruptions to recruitment, consenting, and DNA sampling during that month, particularly in the second half of that month. The Project leadership enabled a plan to initiate remote recruitment and consenting starting in April of 2020. A workflow for remote DNA sample collection took longer to establish and was not fully approved, tested, and operationalized until October of 2020. In **Table 1** below, there is a side-by-side description of the major features of the pre-pandemic face-to-face workflow (July 2019 to March 2020) and the pandemic remote workflows (April 2020 to June 2022). It is noted that the face-to-face workflows with appropriate personal protective equipment continued at a low level during the pandemic where possible (e.g., hospitalized volunteers). The fluctuating month-to-month staffing levels are presented in **Supplementary Table 1**.

**TABLE 1.** Elements of the YGP’s Pre-Pandemic and Pandemic Workflows for Recruitment, Consent and DNA Sample Acquisition.

| MAJOR TASKS | DISCRETE STEPS | Pre-Pandemic Methods<br>(Established<br>June 2019-March 2020) | Pandemic Methods<br>(Added April 2020-June 2022) |
| --- | --- | --- | --- |
| RECRUITMENT | Project Promotion | Consenters setup recruitment tables in both health system and community sites.<br>Consenters distributed project informational documents to interested individuals.<br>Targeted recruitment and consenting within select clinical programs. | YGP leadership and YNHH physicians collaborated to identify groups of patients for outreach.<br>Consenters contacted potential volunteers via electronic messages, phone calls, mailed letters.<br>Public messaging about YGP via website and public announcements. |
|  | Case Identification | Interested volunteers present themselves to the recruiters. | Potential participants responded to contact with expressed interest in project.<br>Self-identified volunteers reached out independently to project. |
| CONSENT | Individualized Informational Exchange | Face-to-face discussion between trained consentor and volunteer. Describing project, review consent document, and answer questions. | Teleconference discussion between trained consentor and volunteer. Describing project, review consent document, and answer questions. |
|  | EHR Registration | Consentor creates an EHR in real time for any previously unregistered volunteers. | Consentor creates an EHR remotely for any previously unregistered volunteers. |
|  | Informed Consent | Consentor has volunteer electronically sign consent document on the computer tablet. | Consentor documents volunteer's verbal consent to YGP during tele-video consent meeting. |
| DNA SAMPLE ACQUISITION | EHR Clinical Order Placement | Signed consent triggers a clinical order for blood draw for whole exome sequence and SNP array. | Verbal consent triggers a clinical order for saliva collection for whole exome sequence and SNP array. |
|  | Volunteer Sample Collection Method | Volunteer presents to any one of the health system's 500 phlebotomy stations for peripheral blood draw to fulfill clinical order in EHR.<br>Volunteer opts for phlebotomy visit as either for project sample alone, or in conjunction with other clinical sample collection. | Saliva collection kit shipped to volunteer's home.<br>Volunteer teleconferences with member of consent team for "coached" sample collection.<br>Volunteer ships sample to YGP office. |

The project staff were able to work from home for most aspects of their jobs during the pandemic period. The exception to that was the need for at least one onsite staff member to process the saliva collection kits that arrived by courier. The processing included accessioning the samples that arrived, and then sending the samples to the hospital’s clinical laboratory for DNA isolation, sequencing, and genotyping.

All volunteers had a YNHH electronic health record (EHR), and this allowed for a de-identified tabulation of data points of interest in **Table 2** including: sex, race, and ethnicity. In addition, whether the volunteer was an established patient within the YNHH or if volunteering for the project prompted a new registration to the health system, as well as whether the volunteer resided within the state of Connecticut or outside of Connecticut, and whether the enrollment was face-to-face or remote.

**TABLE 2.** De-identified Demographic and Related Data from Electronic Health Record (EHR) of the YGP Volunteers over the period of this report.

|  | PRE-PANDEMIC<br>(9 months) | PANDEMIC PERIOD<br>(27 months) |  |  | TOTAL |
| --- | --- | --- | --- | --- | --- |
|  | Face to Face | Face to Face | Remote | Combined |  |
| **Total** | 1950 | 247 | 2752 | 2999 | 4949 |
| **Sex** |  |  |  |  |  |
| Female | 1416 (72.62%) | 140 (56.68%) | 1638 (59.52%) | 1778 (59.29%) | 3194 (64.54%) |
| Male | 534 (27.38%) | 107 (43.32%) | 1114 (40.48%) | 1221 (40.71%) | 1755 (35.46%) |
| **Race** |  |  |  |  |  |
| American Indian or Alaska Native | 8 (0.41%) | 0 (0.00%) | 11 (0.40%) | 11 (0.37%) | 19 (0.38%) |
| Asian | 72 (3.69%) | 6 (2.43%) | 102 (3.71%) | 108 (3.60%) | 180 (3.64%) |
| Black or African American | 265 (13.59%) | 39 (15.79%) | 239 (8.68%) | 278 (9.27%) | 543 (10.97%) |
| Native Hawaiian or Other Pacific Islander | 5 (0.29%) | 0 (0.00%) | 9 (0.33%) | 9 (0.30%) | 14 (0.28%) |
| White or Caucasian | 1349 (69.18%) | 171 (69.23%) | 1943 (70.60%) | 2114 (70.49%) | 3463 (69.97%) |
| Not Provided | 251 (12.87%) | 31 (12.55%) | 448 (16.28%) | 479 (15.97%) | 730 (14.75%) |
| **Ethnicity** |  |  |  |  |  |
| Hispanic or Latino | 223 (11.44%) | 22 (8.91%) | 261 (9.48%) | 283 (9.44%) | 506 (10.22%) |
| Non-Hispanic | 1624 (83.28%) | 213 (86.23%) | 2201 (79.98%) | 2414 (80.49%) | 4038 (81.59%) |
| Not Provided | 103 (5.28%) | 12 (4.86%) | 290 (10.54%) | 302 (10.07%) | 405 (8.18%) |
| **YNHH EHR Established** |  |  |  |  |  |
| Prior to Volunteering | 1833 (94.00%) | 244 (98.79%) | 2717 (98.73%) | 2961 (98.73%) | 4794 (96.87%) |
| In Conjunction with Volunteering | 117 (6.00%) | 3 (1.21%) | 35 (1.27%) | 38 (1.27%) | 155 (3.13%) |
| **Residence** |  |  |  |  |  |
| Inside Connecticut | 1843 (94.51%) | 221 (89.47%) | 2406 (87.43%) | 2627 (87.60%) | 4470 (90.32%) |
| Outside Connecticut | 107 (5.49%) | 26 (10.53%) | 346 (12.57%) | 372 (12.40%) | 479 (9.68%) |

## RESULTS

### Observed Differences in Pre-pandemic and Pandemic Demographics

We observe several trends within the data displayed in **Table 2** below. There was an increase in the percentage of male volunteers and a decrease in the percentage of Hispanic volunteers during the pandemic. The percentage of African-American volunteers decreased and the percentage of those who did not provide race and ethnicity information increased amongst those joining the project remotely. There was an increased percentage of volunteers who were established YNHH patients as well as an increased percentage of volunteers who resided outside of the state of Connecticut during the pandemic period.

### Variation in Monthly Consenting and DNA Sample Collection During this Study

The total number of volunteers who were consented during the 36 months recorded in this manuscript were 4,949. In the same period, the total number of DNA sample collections was 3,926. There is typically a lag in DNA collection following consenting in projects with this design, however a 20.67% gap appears to be exaggerated due to limitations associated with the pandemic ^**[8]**^.

In **Figure 1A and B**, the month-to-month tallies for consenting and sample collection are presented. This information is presented in tabular form in **Supplemental tables 2 and 3**. Both figures demonstrate a drop-off in volunteer consents and samples starting in March 2020, coinciding with the initiation of pandemic precautions and staff re-assignments in the second half of that month.

**FIGURE 1A AND 1B.**
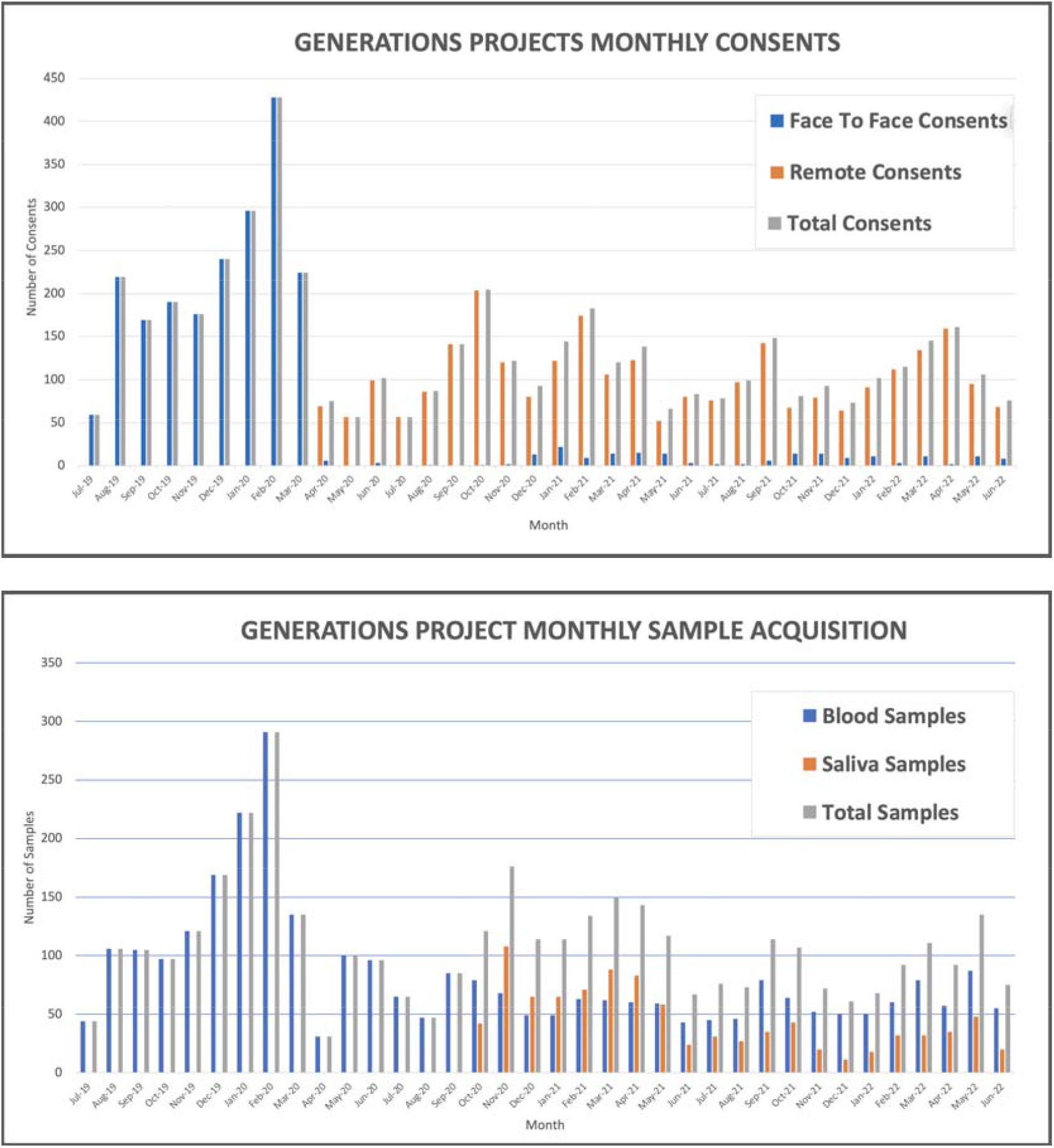
The month-to-month tallies for consenting (1A) and sample acquisition (1B).

As of June 30th 2022 there were N=4949 volunteers enrolled in YGP. Of these, there were N=1,950 (216.7 per month) volunteers consented pre-pandemic, and N=2,999 (111.1 per month) volunteers during pandemic. DNA sample acquisition peaked in the pre-pandemic month of February 2020 with N=291 peripheral blood draws, and in the pandemic period the peak DNA sample acquisition month was November 2020 with N=176 (N=68 peripheral blood draws and N=108 saliva samples).

## DISCUSSION

The project’s trajectory at the end of 2019 and the beginning of 2020 was aligned with the initial project goal of hiring a consent team of 11 FTE responsible for consenting greater than 800 volunteers per month, however that goal was not achieved due to pandemic associated limitations in the period reported here. Nevertheless, the YGP did recruit and consent nearly 3,000 volunteers during the 27 months of COVID restrictions reported here, making it the most successful clinical research project recruitment at Yale during the pandemic.

The observed differences in the volunteer demographics in the pre-pandemic and the pandemic phases are believed to be multi-factorial, and they offer insights into ways to craft a post-pandemic strategy that will allow the project to meet recruitment goals, including increased diversity and inclusion. The project plan is to renew face-to-face recruitment and consenting as part of an effort to increase the number of African American and Hispanic volunteers, while maintaining the remote consenting efforts that increase the geographic reach of the project.

Currently in Connecticut, most of the pandemic related obstacles to YGP recruitment, consenting, and DNA sample acquisition have been resolved. Like other groups around the world, the YGP leadership plans to use the data obtained in the 36-month period described here to inform optimized hybrid workflows for future work ^**[9]**^.

## Data Availability

All data produced in the present work are contained in the manuscript.

## DECLARATIONS

## Acknowledgments

The authors of this manuscript would like to acknowledge Richard Hintz for his work with de-identified database searches.

## Authors’ contributions

Contributed to the conception and design of the study: AM Derry, Y Strong, KM Anastasio MF Murray, Contributions to the acquisition, analysis, or interpretation of data for the work: AM Derry, Y Strong, D Schioppo, J Cotter, GM Wilkins, LI Siqueiros, A Ouyang, K Hulseman, J Petrosino, L Liang, M Stevenson, TE Aguilera, AL Soto, K Meurer, AL Herman, I Cohen, GJ Falcone, EE Longbrake, CI Ochoa Chaar, KM Anastasio, MF Murray

## Availability of data and materials

Not applicable

## Ethical approval and consent to participate

The Yale Institutional Review Board (IRB) approved the protocol and the consent for Yale Generations Project (study HIC#2000024015). All patients were consented via the IRB approved process.

## Consent for publication

Not applicable

## Copyright

© The Author(s) 2021.

