## Supplemental Tables 1, 2, and 3 for "Recruitment, Consent and DNA Sample Acquisition in a U.S. Precision Health Cohort During the COVID-19 Pandemic"

**Supplemental Table 1** – The month-to-month size of the Yale Generations Project Recruitment and Consenting Workforce

| **Month** | **Full Time Equivalents** | **Part Time Equivalents** | **Total** |
| --- | --- | --- | --- |
| **July 2019** | 3.0 | -- | **3.0** |
| **August 2019** | 3.0 | -- | **3.0** |
| **September 2019** | 3.0 | -- | **3.0** |
| **October 2019** | 5.0 | -- | **5.0** |
| **November 2019** | 9.0 | -- | **9.0** |
| **December 2019** | 9.0 | -- | **9.0** |
| **January 2020** | 8.0 | -- | **8.0** |
| **February 2020** | 8.0 | -- | **8.0** |
| **March 2020** | 9.0 | -- | **9.0** |
| **April 2020** | 9.0 | -- | **9.0** |
| **May 2020** | 9.0 | -- | **9.0** |
| **June 2020** | 9.0 | -- | **9.0** |
| **July 2020** | 6.0 | -- | **6.0** |
| **August 2020** | 5.0 | -- | **5.0** |
| **September 2020** | 5.0 | -- | **5.0** |
| **October 2020** | 5.0 | -- | **5.0** |
| **November 2020** | 5.0 | -- | **5.0** |
| **December 2020** | 5.0 | -- | **5.0** |
| **January 2021** | 5.0 | -- | **5.0** |
| **February 2021** | 5.0 | -- | **5.0** |
| **March 2021** | 4.0 | -- | **4.0** |
| **April 2021** | 3.0 | 0.5 | **3.5** |
| **May 2021** | 3.0 | 0.5 | **3.5** |
| **June 2021** | 4.0 | 0.5 | **4.5** |
| **July 2021** | 4.0 | 0.5 | **4.5** |
| **August 2021** | 3.0 | 0.5 | **3.5** |
| **September 2021** | 3.0 | 0.5 | **3.5** |
| **October 2021** | 1.0 | 0.5 | **1.5** |
| **November 2021** | 1.0 | 0.5 | **1.5** |
| **December 2021** | 1.0 | 0.5 | **1.5** |
| **January 2022** | 2.0 | 0.5 | **2.5** |
| **February 2022** | 2.0 | 0.5 | **2.5** |
| **March 2022** | 2.0 | 0.5 | **2.5** |
| **April 2022** | 2.0 | 0.5 | **2.5** |
| **May 2022** | 2.0 | 0.5 | **2.5** |
| **June 2022** | 2.0 | 0.5 | **2.5** |

**Supplemental Table 2** – The month-to-month consenting for the Yale Generations Project this is the data used to construct **Figure 1A**

| **Consent Year/Month** | **Remote** | **Face-to-Face** | **Total** |
| --- | --- | --- | --- |
| ** July 2019 | 0 | 59 | 59 |
| August 2019 | 0 | 219 | 219 |
| September 2019 | 0 | 169 | 169 |
| October 2019 | 0 | 190 | 190 |
| November 2019 | 0 | 176 | 176 |
| December 2019 | 0 | 240 | 240 |
| January 2020 | 0 | 296 | 296 |
| February 2020 | 0 | 428 | 428 |
| March 2020 | 0 | 224 | 224 |
| April 2020 | 69 | 6 | 75 |
| May 2020 | 56 | 0 | 56 |
| June 2020 | 99 | 3 | 102 |
| July 2020 | 56 | 0 | 56 |
| August 2020 | 86 | 1 | 87 |
| September 2020 | 141 | 0 | 141 |
| October 2020 | 204 | 1 | 205 |
| November 2020 | 120 | 2 | 122 |
| December 2020 | 80 | 13 | 93 |
| January 2021 | 122 | 22 | 144 |
| February 2021 | 174 | 9 | 183 |
| March 2021 | 106 | 14 | 120 |
| April 2021 | 123 | 15 | 138 |
| May 2021 | 52 | 14 | 66 |
| June 2021 | 80 | 3 | 83 |
| July 2021 | 76 | 2 | 78 |
| August 2021 | 97 | 2 | 99 |
| September 2021 | 142 | 6 | 148 |
| October 2021 | 67 | 14 | 81 |
| November 2021 | 79 | 14 | 93 |
| December 2021 | 64 | 9 | 73 |
| January 2022 | 91 | 11 | 102 |
| February 2022 | 112 | 3 | 115 |
| March 2022 | 134 | 11 | 145 |
| April 2022 | 159 | 2 | 161 |
| May 2022 | 95 | 11 | 106 |
| June 2022 | 68 | 8 | 76 |
| **TOTALS** | 2752 | 2197 | 4949 |

** July 2019 numbers includes 12 volunteers consented at the end of June 2019

**Supplemental Table 3** – The month-to-month sample acquisition for the Yale Generations Project this is the data used to construct **Figure 1B**

| **Date** | **Blood Samples** | **Saliva Samples** | **Total Samples** |
| --- | --- | --- | --- |
| ** July 2019 | 44 | 0 | 44 |
| August 2019 | 106 | 0 | 106 |
| September 2019 | 105 | 0 | 105 |
| October 2019 | 97 | 0 | 97 |
| November 2019 | 121 | 0 | 121 |
| December 2019 | 169 | 0 | 169 |
| January 2020 | 222 | 0 | 222 |
| February 2020 | 291 | 0 | 291 |
| March 2020 | 135 | 0 | 135 |
| April 2020 | 31 | 0 | 31 |
| May 2020 | 100 | 0 | 100 |
| June 2020 | 96 | 0 | 96 |
| July 2020 | 65 | 0 | 65 |
| August 2020 | 47 | 0 | 47 |
| September 2020 | 85 | 0 | 85 |
| October 2020 | 79 | 42 | 121 |
| November 2020 | 68 | 108 | 176 |
| December 2020 | 49 | 65 | 114 |
| January 2021 | 49 | 65 | 114 |
| February 2021 | 63 | 71 | 134 |
| March 2021 | 62 | 88 | 150 |
| April 2021 | 60 | 83 | 143 |
| May 2021 | 59 | 58 | 117 |
| June 2021 | 43 | 24 | 67 |
| July 2021 | 45 | 31 | 76 |
| August 2021 | 46 | 27 | 73 |
| September 2021 | 79 | 35 | 114 |
| October 2021 | 64 | 43 | 107 |
| November 2021 | 52 | 20 | 72 |
| December 2021 | 50 | 11 | 61 |
| January 2022 | 50 | 18 | 68 |
| February 2022 | 60 | 32 | 92 |
| March 2022 | 79 | 32 | 111 |
| April 2022 | 57 | 35 | 92 |
| May 2022 | 87 | 48 | 135 |
| June 2022 | 55 | 20 | 75 |
| **TOTALS** | 2970 | 956 | 3926 |

** July 2019 numbers includes 12 samples collected at the end of June 2019
